# Development and validation of blood-based prognostic biomarkers for severity of COVID disease outcome using EpiSwitch 3D genomic regulatory immuno-genetic profiling

**DOI:** 10.1101/2021.06.21.21259145

**Authors:** Ewan Hunter, Christina Koutsothanasi, Adam Wilson, Francisco C. Santos, Matthew Salter, Jurjen W. Westra, Ryan Powell, Ann Dring, Paulina Brajer, Benedict Egan, Matthew Parnall, Catriona Williams, Aemilia Katzinski, Thomas Lavin, Aroul Ramadass, William Messer, Amanda Brunton, Zoe Lyski, Rama Vancheeswaran, Andrew Barlow, Dmitri Pchejetski, Peter A. Robbins, Jane Mellor, Alexandre Akoulitchev

## Abstract

The COVID-19 pandemic has raised several global public health challenges to which the international medical community have responded. Diagnostic testing and the development of vaccines against the SARS-CoV-2 virus have made remarkable progress to date. As the population is now faced with the complex lifestyle and medical decisions that come with living in a pandemic, a forward-looking understanding of how a COVID-19 diagnosis may affect the health of an individual represents a pressing need. Previously we used whole genome microarray to identify 200 3D genomic marker leads that could predict mild or severe COVID-19 disease outcomes from blood samples in a multinational cohort of COVID-19 patients. Here, we focus on the development and validation of a qPCR assay to accurately predict severe COVID-19 disease requiring intensive care unit (ICU) support and/or mechanical ventilation. From 200 original biomarker leads we established a classification model containing six markers. The markers were qualified and validated on 38 COVID-19 patients from an independent cohort. Overall, the six-marker model obtained a positive predictive value of 93% and balanced accuracy of 88% across 116 patients for the prognosis of COVID-19 severity requiring ICU care/ventilation support. The six-marker signature identifies individuals at the highest risk of developing severe complications in COVID-19 with high predictive accuracy and can assist in patient prognosis and clinical management decisions.

## Background

The COVID-19 outbreak, which the World Health Organization (WHO) declared a pandemic in March 2020, represents one of the greatest global health crises the world has faced in recent history [1]. In addition to the estimated 130+ million people that have been infected with the SARS-CoV-2 virus to date and the more than 3 million deaths attributed to COVID-19 related causes; the pandemic has placed tremendous strain on healthcare systems, caused devastating mental health crises, and tested global economic resiliency [2, 3]. In response to the COVID-19 crisis, the global research and medical communities responded quickly with the rapid development and deployment of diagnostic tests to identify individuals who are currently infected or have previously been infected with the SARS-CoV-2 virus. As of March 2021, over 250 different rapid antigen or molecular tests to detect SARS-CoV-2 have received Emergency Use Authorization by the US FDA alone [4, 5]. For several reasons, including the early immediate unmet need to identify SARS-CoV-2 positive individuals and the lack of availability of real-world data on how different individuals respond to the virus, research on features that are predictive for different manifestations of COVID-19 have lagged behind the development of diagnostic testing. The few predictive measures that have been assessed suffer from low certainty, high bias, and insufficient predictive accuracy [6]. As clinicians have gained experience in treating COVID-19 patients, one of the more striking observations has been the wide range of clinical manifestations of the disease in different individuals; many of which could not be predicted by basic clinical parameters such as age, gender, or pre-existing conditions/co-morbidities alone [7–9]. It is now clear that the variability in host response, rather than viral genetics or load, is the primary determinant of the wide range of observed disease manifestations.

While diagnostic testing is now routine, there remains a pressing need to develop molecular readouts that can reveal individual susceptibility to developing severe COVID-19 symptoms in response to the SARS-CoV-2 virus with high predictive accuracy to support clinical decision making and patient management [10]. Specifically, there is a need to develop accurate, non-invasive tests to identify prognostic profiles for clinical cases where patients develop hyperinflammation, do not respond to the standard of care, and progress to needing ICU-level support and/or mechanical ventilation in accordance with the WHO severity definitions [10]. Individualized knowledge of disease severity risk can help people take appropriate precautions in their daily lives, understand how a COVID-19 diagnosis may impact a pre-existing medical condition, anticipate and plan for medical treatment if they become infected, and decrease their personal risk of contracting severe symptoms by managing risk factors. All of these are critical tools for managing the global health crisis, even in the face of the recent development of COVID-19 vaccines. The first vaccines against the SARS-CoV-2 virus became available in the late 2020 and represented a major step forward in containing the spread of the virus and its myriad social tolls [11–13]. However, several factors may limit the effectiveness of universal vaccination: 1) vaccine shortages and limited national vaccination rates 2) contraindications in certain patient populations (e.g. children under 18, individuals who have certain health conditions, such as recent infections and anaphylaxis) 3) individual decisions to forego vaccination and 4) reductions in vaccine efficacy in newly emerging SARS-CoV-2 variants as demonstrated recently in a Chilean population with persistently high infection rates despite the availability of the CoronaVac vaccine and >40% national vaccination rates [14]. As such, the ability for individuals and their physicians to understand the risk of developing severe symptoms if infected with the SARS-CoV-2 virus remains a critical unmet healthcare need.

Recently, we reported on the use of *EpiSwitch*®, a chromosome conformation capture (3C) methodology that has been reduced to practice for the discovery of blood-based 3D genomic biomarkers and patient stratification in a variety of immune-related diseases, to identify prognostic profiles that could serve as a potential blood-based molecular classifier for the prediction of COVID-19 disease severity [15–22]. Specifically, we used *EpiSwitch* Explorer 3D whole genome arrays to identify changes at immune-related loci in blood samples from a multi-national cohort of COVID-19 patients. We identified 200 array-based 3D genomic marker leads that could differentially predict mild or severe COVID-19 disease severity, with a high degree of sensitivity and specificity. The proteins encoded at the locations of the 3D genomic marker leads were involved in biological pathways with direct relevance to immune system function including T-cell signalling, macrophage-stimulating protein (MSP)-RON signalling, and calcium signalling [15]. The aim of this study was an extension of our previous work, using the established *EpiSwitch* platform to translate the array-derived 3D genomic biomarkers into a qPCR-based testing format, refine and reduce the biomarker panel, build a classification model and perform validation on independent COVID-19 patient cohorts. Here we report on the development of a 3D genomic biomarker panel with clinical utility in predicting disease severity in COVID-19.

## Materials and Methods

### Patient characteristics

Clinical whole blood samples from consented patients were supplied from Boca Biolistics LLC (FL, USA) and Reprocell USA Inc. (MD, USA). A total of 116 patients from three sample cohorts were used in this study, comprising a multinational set of COVID-19 cases from the United States, Peru, and the Dominican Republic. Patient annotations are listed in (**Supplemental Tables 1,2**). In line with WHO guidelines, the patient annotations provided were used to classify the severe outcome group on the basis of a confirmed admission to the Intensive Care Unit (ICU) and/or advanced clinical interventions such as mechanical ventilation [10]. Patients that required a lower level of clinical care, such as administration of supplemental oxygen only, were classified as the mild outcome group. All samples were collected within 72 hours of a patient being admitted to a hospital for treatment of a PCR-confirmed COVID infection. The age of the patients ranged from 28 to 92, with median of 64.5 years (**Supplemental Tables 1, 2**).

### Preparation of 3D genomic templates

*EpiSwitch^®^* 3C libraries, with chromosome conformation analytes converted to sequence based tags, were prepared from frozen whole blood samples using *EpiSwitch^®^* protocols following the manufacturer’s instructions (Oxford BioDynamics Plc) [15]. All samples were processed under biological containment level CL2+. Initial sample processing was performed manually in a Category 3 microbial safety cabinet with the remainder performed on the Freedom EVO 200 robotic platform (Tecan Group Ltd). Briefly, 50 μL of whole blood sample was diluted and fixed with a formaldehyde containing *EpiSwitch* buffer. Density cushion centrifugation was used to purify intact nuclei. Following a short detergent-based step to permeabilise the nuclei, restriction enzyme digestion and proximity ligation were used to generate the 3C libraries. Samples were centrifuged to pellet the intact nuclei before purification with an adapted protocol from the QIAmp DNA FFPE Tissue kit (Qiagen) Eluting in 1x TE buffer pH7.5. 3C libraries were quantified using the Quant-iT™ Picogreen dsDNA Assay kit (Invitrogen) and normalised to 5 ng/μl prior to interrogation by PCR.

### Translation of array-based 3D genomic markers to PCR readouts

The top array-derived markers identified in our previous study were interrogated using OBD’s proprietary primer design software package to identify genomic positions suitable for a hydrolysis probe based real time PCR (RT-PCR) assay [15]. Briefly, the top array-derived markers associated with prognostic potential to differentiate between mild and severe COVID disease outcomes were filtered on fold change and adjusted p value. PCR primer probes were ordered from Eurofins genomics as salt-free primers. The probes were designed with a 5’ FAM fluorophore, 3’ IABkFQ quencher and an additional internal ZEN quencher and ordered from iDT (integrated DNA Technologies) [23]. Each assay was optimised using a temperature gradient PCR with an annealing temperature range from 58-68° C. Individual PCR assays were tested across the temperature gradient alongside negative controls including soluble and unstructured commercial TaqMan human genomic DNA control (Life Technologies) and used a TE buffer only negative control. Assay performance was assessed based on Cq values and reliability of detection and efficiency based on the slope of the individual amplification curves. Assays that passed the quality criteria and presented with reliable detection differences between the pooled samples associated with Severe and Mild COVID disease outcomes were used to screen individual patient samples.

### EpiSwitch PCR

Each patient sample was interrogated using RT-PCR in triplicate. Each reaction consisted of 50 ng of EpiSwitch library template, 250 mM of each of the primers, 200 mM of the hydrolysis probe and a final 1X Kapa Probe Force Universal (Roche) concentration in a final 25 μl volume. The PCR cycling and data collection was performed using a CFX96 Touch Real-Time PCR detection system (Bio-Rad). The annealing temperature of each assay was changed to the optimum temperature identified in the temperature gradients performed during translation for each assay. Otherwise, the same cycling conditions were used: 98°C for 3 minutes followed by 45 cycles of 95°C for 10 seconds and 20 seconds at the identified optimum annealing temperature. The individual well Cq values were exported from the CFX manager software after baseline and threshold value checks. All Cq values obtained for individual samples and markers are available online [https://github.com/oxfordBiodynamics/medrxiv/tree/main/CST%20publication]

A total of 21 3D genomic markers that passed the translation phase were screened on 78 individual samples from the Training cohort. A marker reduction step based on statistical criteria were used to identify the top six discriminating markers which were used to screen the remaining set of 38 samples in the Test cohort.

### Genomic mapping

The 21 3D genomic markers from the statistically filtered list with the greatest and lowest abundance scores were selected for genome mapping. Mapping was carried out using Bedtools *closest* function for the 3 closest protein coding loci (Gencode v33). All markers were visualized using the *EpiSwitch* Data Portal.

### Statistical analysis

The 21 markers screened on 78 individual patient samples were subject to permutated logistic modelling with bootstrapping for 500 data splits and non-parametric Rank Product analysis (EpiSwitch® RankProd R library). Two machine learning procedures (eXtreme Gradient Boosting: XGBoost and CatBoost) were used to further reduce the feature pool and identify the most predictive/prognostic, 3D genomic markers. The resulting markers were then used to build the final classifying models using CatBoost and XGBoost. All analysis was performed using R statistical language with Caret, XGBoost, SHAPforxgboost and CatBoost libraries.

### Biological network/pathway analysis

Pathway enrichment analysis was performed using the Reactome Pathway Browser [24]. Protein interaction networks were generated using the Search Tool for the Retrieval of Interacting proteins (STRING) database [24, 25].

## Results

### Identification of the top prognostic 3D genomic markers for severe COVID-19 disease outcomes

In this study we employed a sequential stepwise strategy to identify a minimal set of biomarkers that were predictive of COVID-19 disease severity (**Figure 1**). Starting with an initial set of 200 array-derived 3D genomic marker leads identified in our previously published work, here we used independent sets of 116 whole blood samples collected at the time of confirmed COVID-19 infection from patients in the United States, Peru, and the Dominican Republic to refine and test the prognostic 3D genomic biomarker panel. Whole blood samples were procured for a Training cohort used to build and refine the classifier model, and Test cohort to assess the predictive performance of the model. Clinical characteristics of the patients are shown in **Table 1 and Supplemental Tables 1, 2**. To translate the *EpiSwitch* Explorer Array markers to a PCR-detectable assay, primers to detect individual 3D genomic markers were generated and validated (Materials & Methods). Using a 78 sample Training set of COVID-19 patient whole blood samples, feature reduction using machine learning methods on the initial pool of 200 3D genomic biomarkers identified 21 markers with predictive power to differentiate between COVID-19 patients that required a high degree of medical disease management (e.g. admission to the intensive care unit (ICU), mechanical ventilation) and those that were hospitalized but required less interventional care and support (Supplemental **Table 1, for Cq data see online link –** https://github.com/oxfordBiodynamics/medrxiv/tree/main/CST%20publication).

**Figure 1.**
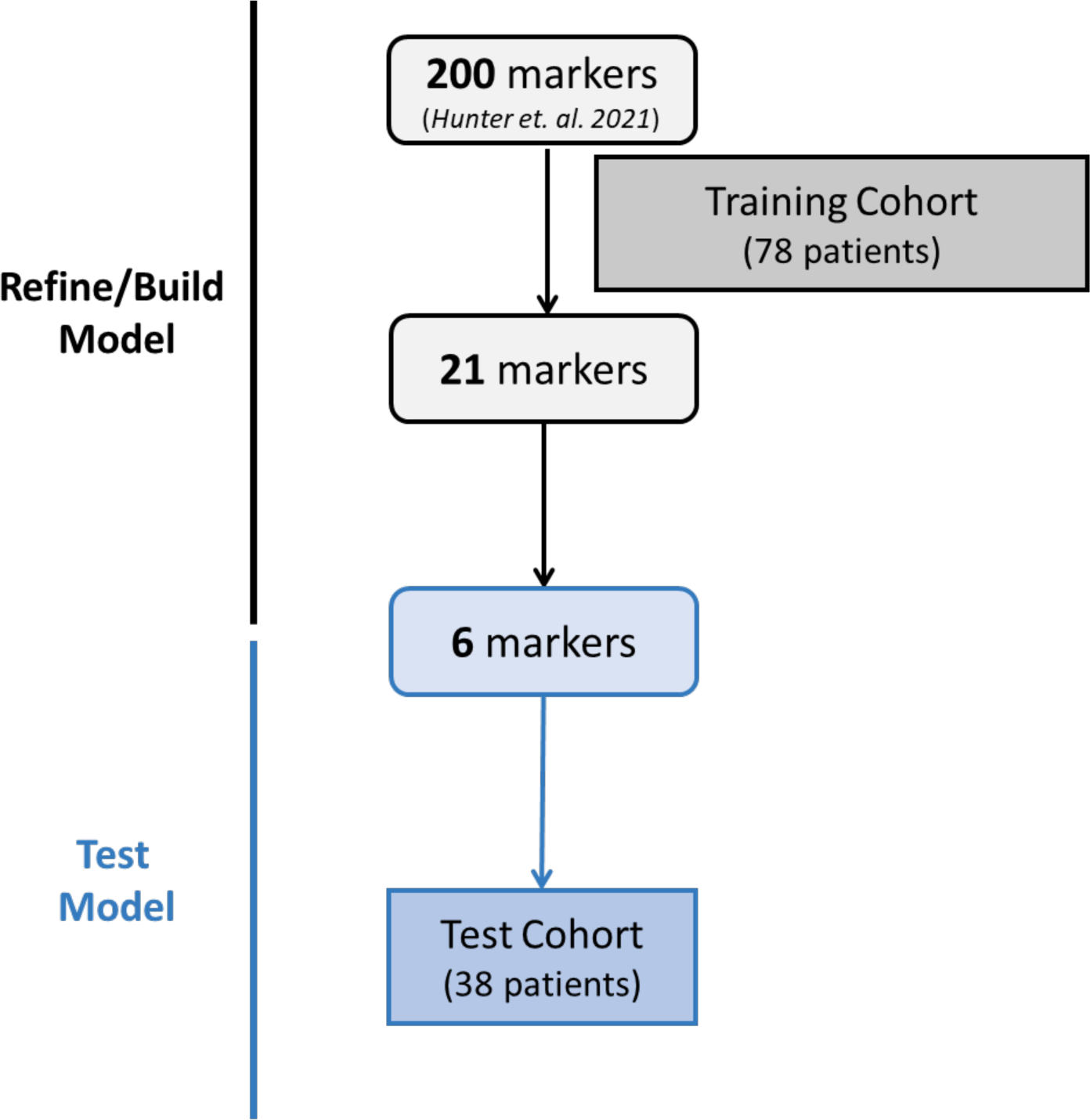
Simplified workflow used to develop and test the prognostic 3D genomic classifier model for prediction of COVID-19 disease severity. Starting from a list of 200 3D genomic markers identified by Hunter et al. 2021, we used a sequential, stepwise approach employing a 78-patient Training cohort to refine the marker set and build a predictive classifier model containing six 3D genomic markers. The 6-marker model/assay was tested on an independent Test cohort of 38 COVID-19 patient blood samples.

**Table 1.** Summary of clinical characteristics by cohort for COVID-19 patient samples used in this study. Clinical features of the 116 COVID-19 patient samples used in this study by cohort. SD: Standard deviation; PEC: pre- existing condition; Hosp.: Hospitalized; ICU: Intensive Care Unit; SO: Received supplemental oxygen; Vent.: Received mechanical ventilation.

| **Cohort** | **N** | **% male** | **% female** | **Age (mean)** | **Age (SD<sup>a</sup>)** | **% w/PEC<sup>b</sup>** | **% Hosp.<sup>c</sup>** | **% ICU<sup>d</sup>** | **% SO<sup>e</sup>** | **% Vent.<sup>f</sup>** |
| --- | --- | --- | --- | --- | --- | --- | --- | --- | --- | --- |
| **Training** | 78 | 64 | 36 | 64.8 | 14.5 | 76 | 46 | 54 | 37 | 60 |
| **Test** | 38 | 79 | 21 | 65.1 | 16.2 | 90 | 53 | 47 | 29 | 74 |

<sup>a</sup> Standard deviation
<sup>b</sup> pre-existing condition
<sup>c</sup> Hospitalized
<sup>d</sup> Intensive Care Unit
<sup>e</sup> Received supplemental oxygen
<sup>f</sup> Received mechanical ventilation

**Table 2.**
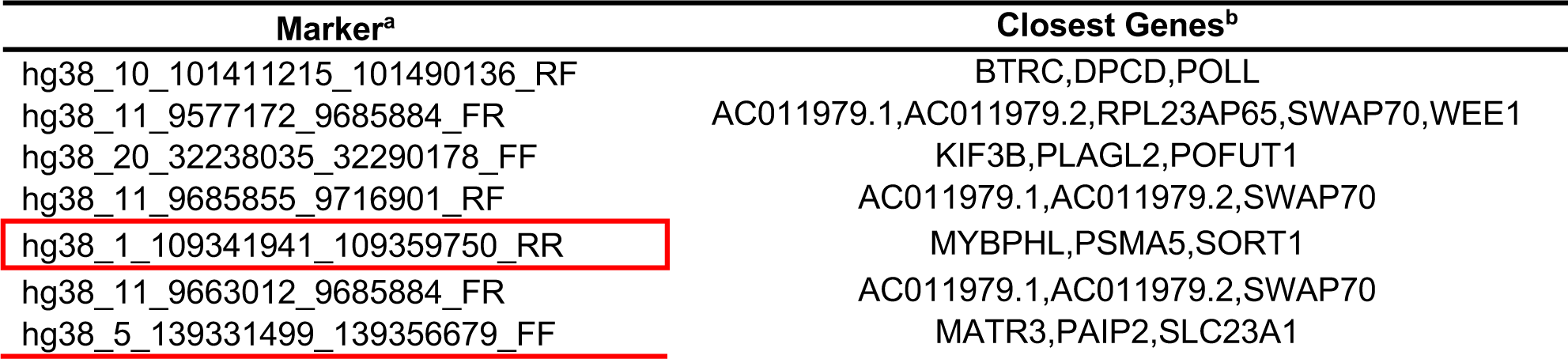
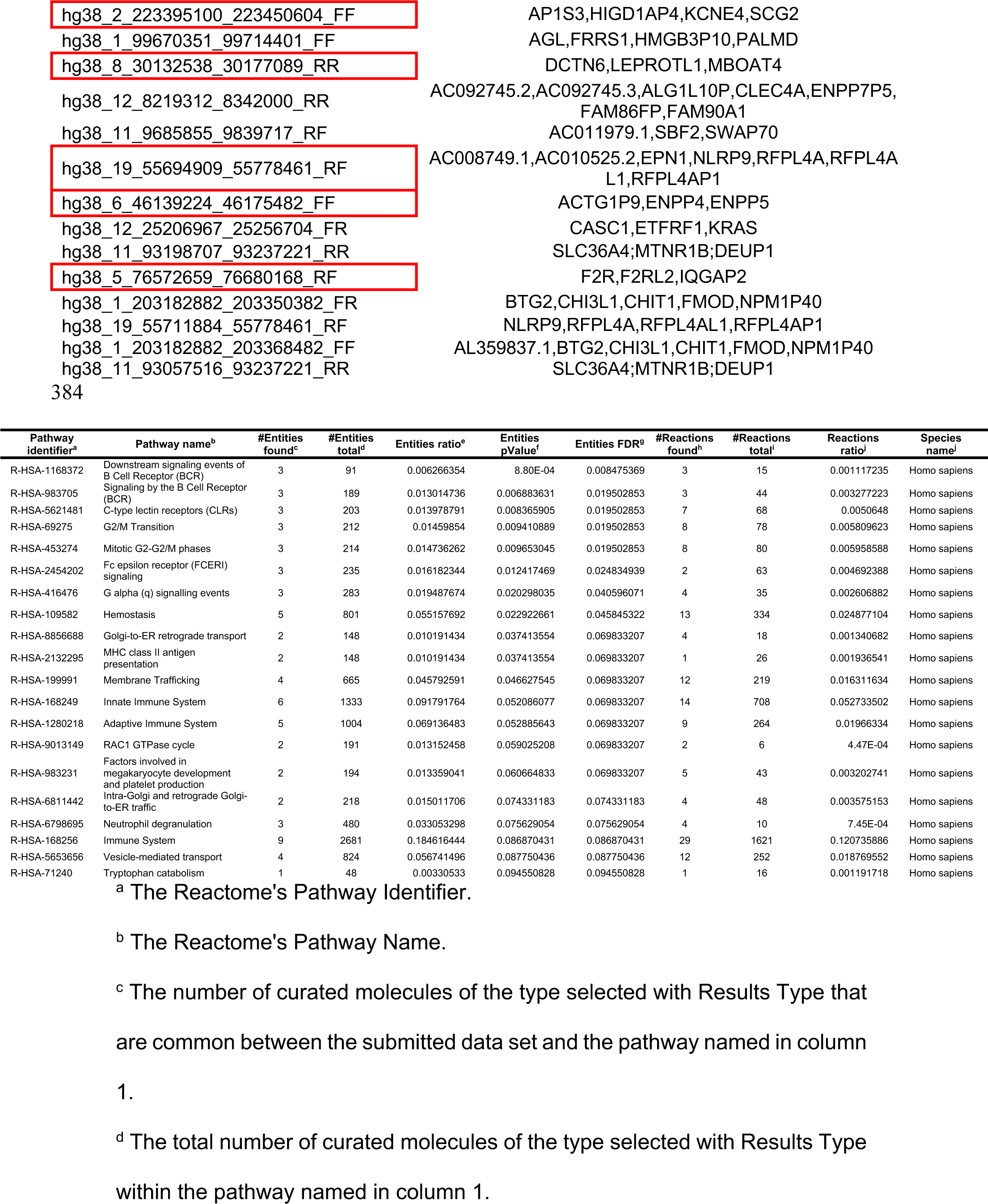
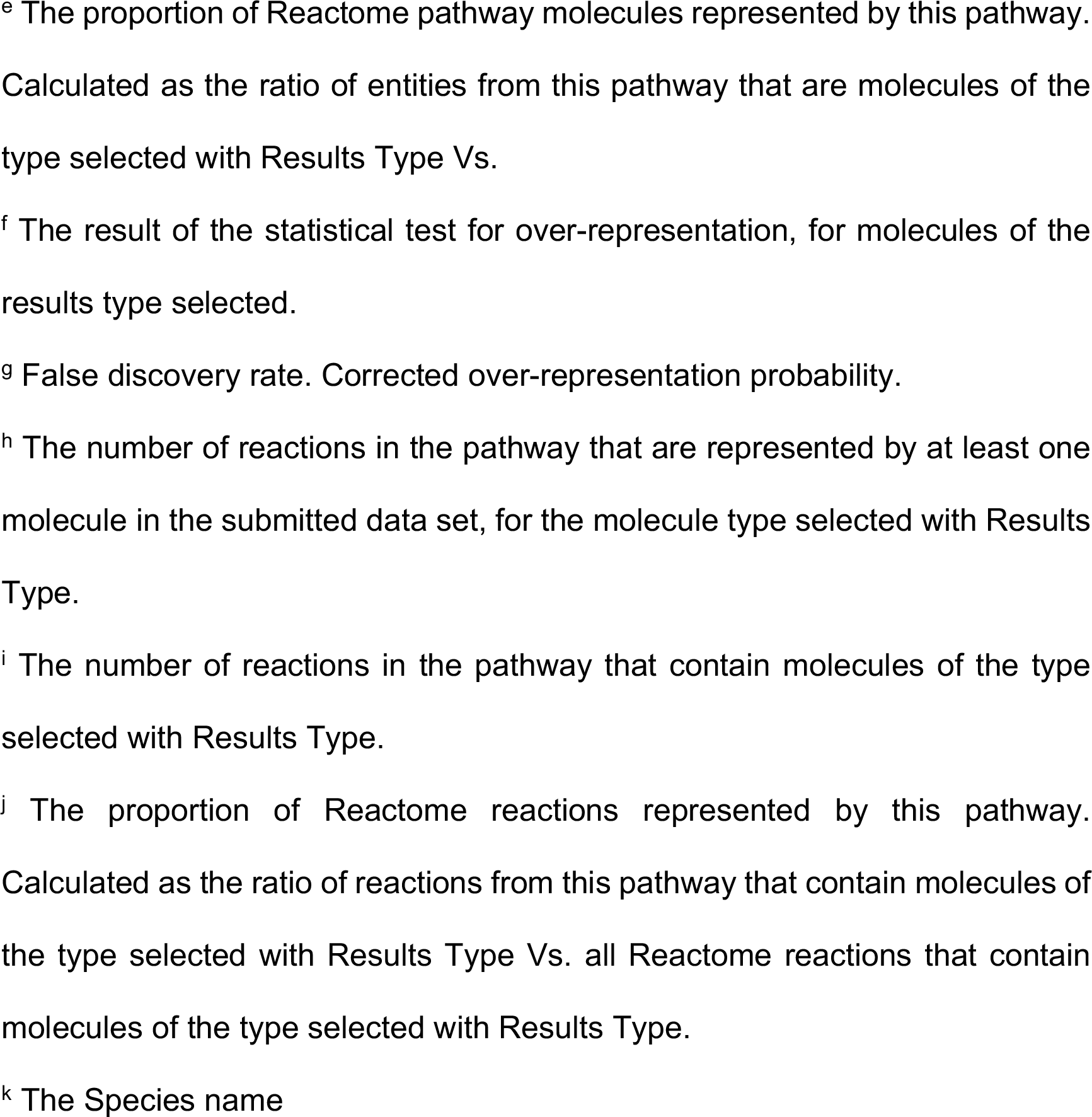
List of the top 21 3D genomic markers for prediction of COVID-19 disease severity. List of the top 21 markers with predictive ability for COVID-19 severity. Markers are listed by the OBD internal ID. The six markers in the final 3D genomic panel are outlined in red. The closest protein-coding genes near the 3D genomic markers are also listed (Closest Genes).

We found the top 21 markers to be non-randomly distributed throughout the human genome, with notable enrichment on larger chromosomes and a hotspots on chromosome 11 (**Figure 2A**). Four out of the 21 markers associated with ICU outcomes occurred within an approximately 265 kb region on the p-arm of chromosome 11 containing the switching B cell complex subunit SWAP70 (SWAP70) locus (**Figure 2B**). While some of the 3D genomic markers were localized within protein coding regions of single genes (**Figure 2C**), other markers spanned the protein coding regions of multiple genes (**Figure 2B**). Pathway enrichment for genes contained within the 21 3D genomic markers revealed the top two pathways to be related to downstream signalling mediated by B-cell receptor activation (**Table 3**). Importantly, genomic loci encoding proteins involved in hemostasis/clotting were also enriched (**Figure 3, Table 3)**. The 21 3D genomic markers were further refined to a set of 6 markers (**Table 4** with predictive ability for COVID severity and applied to an independent Test cohort (Supplemental **Table 2**).

**Figure 2.**
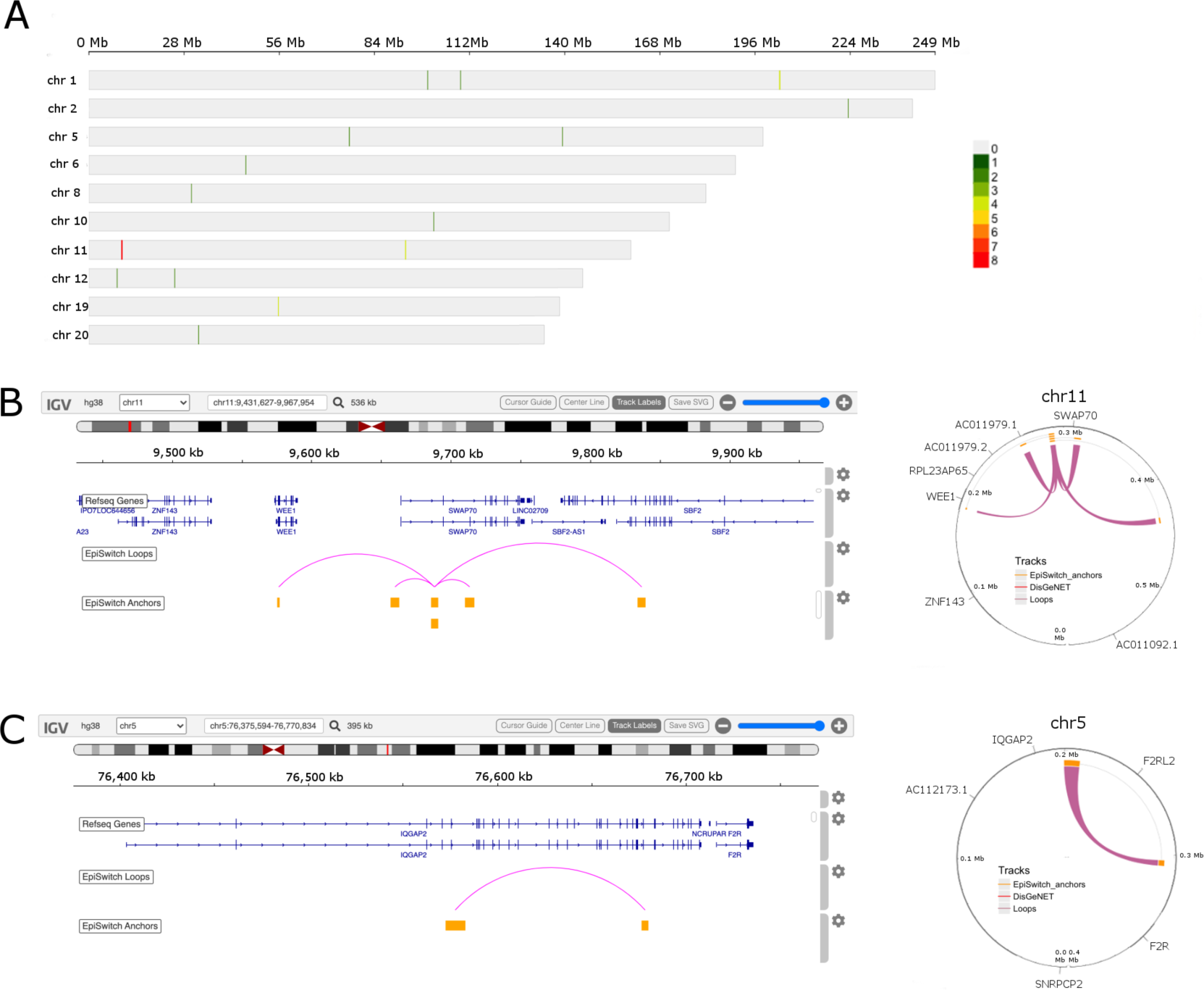
Genomic detail view of top prognostic 3D genomic biomarkers. **A.** Genomic locations and distribution of the top 21 3D genomic markers for Hospitalized and ICU clinical outcomes. Individual human chromosomes where the top 21 markers were found are shown on the y-axis. The heatmap shows the number of markers within a 0.3Mb genomic window with green representing a low density of markers and red indicating a high density of markers. A region on chromosome 11 shows a high density of markers associated with ICU outcomes. **B.** Linear and circos plot views of a ∼500 kb region of chromosome 11 containing the SWAP70 locus showing the genomic location for four markers. **C**. Linear and circos views of a ∼400 kb region of chromosome 5 containing the IQGAP2 and F2RL2 loci showing the genomic location for one of the markers in the final 6-marker set.

**Figure 3.**
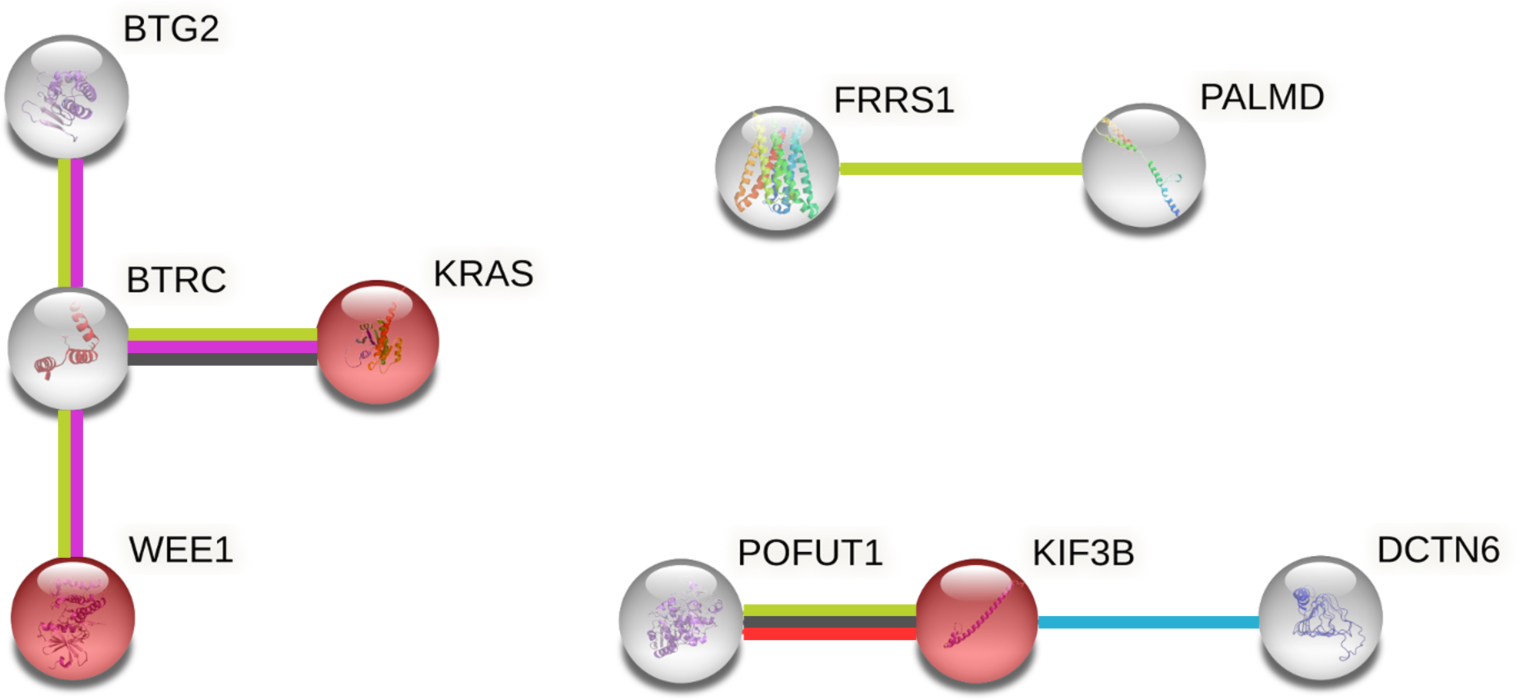
STRING network for top 3D genomic markers. Protein-protein interaction network for proteins encoded by genes spanning the top 21 3D genomic marker set. Edges are colored by protein-protein association type (blue = known interactions from curated databases, magenta = known interactions from experiments, light green = interactions derived from literature text mining, orange = gene fusions, black = association by co-expression). Nodes highlighted in red are associated with the Reactome ‘Hemostasis’ pathway (HSA-109582).

**Table 4.**
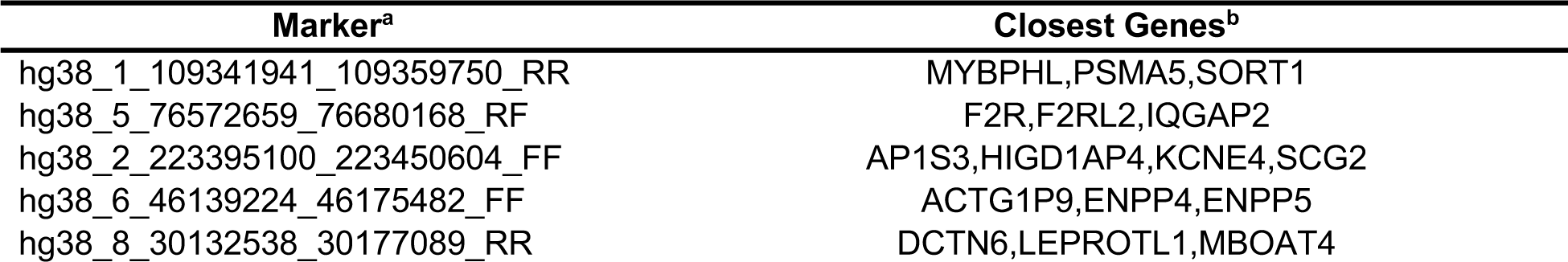
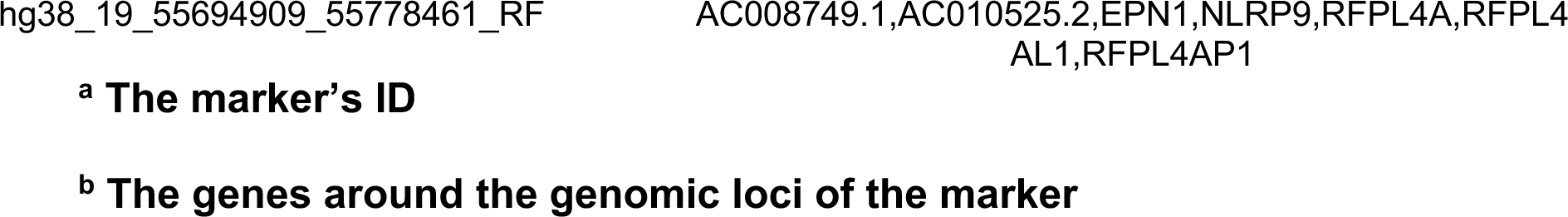
List of the top 6 3D genomic markers for prediction of COVID-19 disease severity. List of the 6 markers that comprise the predictive COVID-19 severity biomarker panel. Markers are listed by the OBD internal ID. The closest protein-coding genes near the 3D genomic markers are also listed in the ‘Closest Genes’ column.

### Testing of the prognostic 3D genomic biomarker panel for severe COVID-19 disease outcomes on independent patient cohorts

To assess the predictive power of the model, the 6-marker 3D genomic panel was validated on an independent (samples that were not used to build and refine the model) Test cohort (**Figure 4, Supplemental Table 2**). Samples were collected upon admission to COVID hospital wards in Peru, the USA, and the Dominican Republic and shipped to OBD’s processing facility in Oxford, UK. The *EpiSwitch* platform read outs for the six-marker classifier model were uploaded to the *EpiSwitch* Analytical Portal for analysis. Classifier calls for high-risk COVID-19 disease outcomes are shown in **Table 5**. Clinical outcomes for the Test cohort included 10 mild cases or 28 severe cases requiring ventilation and/or ICU support. *EpiSwitch* prognostic calls based on the 6-marker model demonstrated performance of 90.9% positive predictive value for high-risk disease outcomes in the Test cohort (**Figure 4A**). Interestingly, two of the mild case patients (COVID 0696 and 0213) (**Table 5**), identified as high risk by the *EpiSwitch* test subsequently died in the hospital within 28 days of admission. This suggests an early, pre-symptomatic detection of a hyperinflammatory state leading to fatal outcomes and is being investigated further. Across all 116 patients used in this study, the test demonstrated positive predictive value for high-risk disease outcomes of 92.9 with, 88% sensitivity, 87% specificity, and a balanced accuracy of 87.9% (**Table 4B and Supplementary Table 3**).

**Figure 4.**
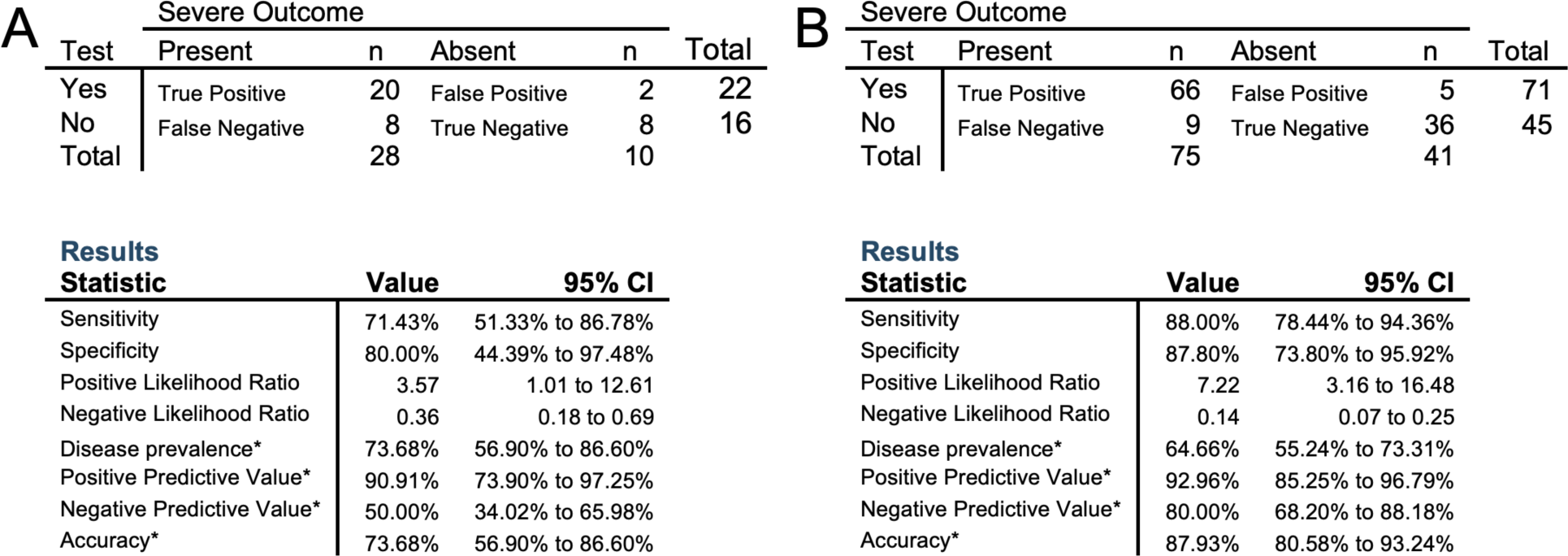
Performance of the prognostic biomarker classifier for calling high risk of severe disease outcome on COVID-19 patient cohorts. (A) Confusion matrix and test performance statistics for the 6-marker classifier on the 38 patients of the Test cohort (B) and the 116 patients in the combined Training and Test cohorts. Note: (*) These values are dependent on disease prevalence

**Table 5.** Prognostic calls for high-risk of severe outcome with the 6-marker *EpiSwitch* classifier model. . Prognostic calls of average and high risk for the 38 patients against disclosed attributes of severity: ventilation and /or ICU support

| Cohort | SampleID | COVID Severity |  | EpiSwitch Prognostic Call for High-Risk |  |  |
| --- | --- | --- | --- | --- | --- | --- |
|  |  | Ventilation | ICU<sup>a</sup> | No | Yes | Final Call |
| Test | COVID0732 | No | No | 0.624535561 | 0.3754644 | No |
| Test | COVID0129 | No | No | 0.989352465 | 0.0106475 | No |
| Test | COVID0636 | No | No | 0.810631394 | 0.1893686 | No |
| Test | COVID0189 | No | No | 0.96364671 | 0.0363533 | No |
| Test | COVID0708 | No | No | 0.918016613 | 0.0819834 | No |
| Test | COVID0117 | No | No | 0.760194659 | 0.2398053 | No |
| Test | COVID0207 | No | No | 0.740656555 | 0.2593434 | No |
| Test | COVID0380 | No | No | 0.990677834 | 0.0093222 | No |
| Test | COVID0696 | No | No | 0.020404769 | 0.9795952 | Yes* |
| Test | COVID0213 | No | No | 0.04568797 | 0.954312 | Yes* |
| Test | COVID0606 | Yes | No | 0.809160769 | 0.1908392 | No |
| Test | COVID0648 | Yes | No | 0.987436414 | 0.0125636 | No |
| Test | COVID0642 | Yes | No | 0.665544152 | 0.3344558 | No |
| Test | COVID0516 | Yes | Yes | 0.601811945 | 0.3981881 | No |
| Test | COVID0564 | Yes | Yes | 0.942398548 | 0.0576015 | No |
| Test | COVID0450 | Yes | Yes | 0.885789573 | 0.1142104 | No |
| Test | COVID0714 | Yes | No | 0.888814926 | 0.1111851 | No |
| Test | COVID0408 | Yes | Yes | 0.700852036 | 0.299148 | No |
| Test | COVID0558 | Yes | Yes | 0.056852765 | 0.9431472 | Yes |
| Test | COVID0540 | Yes | Yes | 0.26985541 | 0.7301446 | Yes |
| Test | COVID0444 | Yes | Yes | 0.012335699 | 0.9876643 | Yes |
| Test | COVID0456 | Yes | Yes | 0.34420839 | 0.6557916 | Yes |
| Test | COVID0468 | Yes | Yes | 0.26985541 | 0.7301446 | Yes |
| Test | COVID0498 | Yes | Yes | 0.045760725 | 0.9542393 | Yes |
| Test | COVID0576 | Yes | Yes | 0.057154838 | 0.9428452 | Yes |
| Test | COVID0504 | Yes | Yes | 0.006351133 | 0.9936489 | Yes |
| Test | COVID0600 | Yes | No | 0.106978044 | 0.893022 | Yes |
| Test | COVID0672 | Yes | No | 0.08792568 | 0.9120743 | Yes |
| Test | COVID0588 | Yes | No | 0.028880829 | 0.9711192 | Yes |
| Test | COVID0654 | Yes | No | 0.029438535 | 0.9705615 | Yes |
| Test | COVID0666 | Yes | No | 0.124919437 | 0.8750806 | Yes |
| Test | COVID0726 | Yes | No | 0.198130682 | 0.8018693 | Yes |
| Test | COVID0474 | Yes | Yes | 0.077650517 | 0.9223495 | Yes |
| Test | COVID0432 | Yes | Yes | 0.145361423 | 0.8546386 | Yes |
| Test | COVID0462 | Yes | Yes | 0.06204395 | 0.9379561 | Yes |
| Test | COVID0510 | Yes | Yes | 0.248548523 | 0.7514515 | Yes |
| Test | COVID0768 | Yes | Yes | 0.377659917 | 0.6223401 | Yes |
| Test | COVID0427 | Yes | Yes | 0.349406302 | 0.6505937 | Yes |

<sup>a</sup> Intensive Care Unit
Note: (\*) these patients were called as high-risk an initially annotated clinically as mild-COVID cases. They subsequently died in the hospital within 28 days of admission

## Discussion

The COVID-19 pandemic will represent a major public health crisis for months to come. As a corollary, there remains a pressing need for prognostic testing that can give individuals a readout of likely disease severity if infected with SARS-CoV2, despite the rigorous social measures put in place to slow the spread of the disease and the recent availability of vaccination options. Despite the development and approval of vaccines by Pfizer-BioNTech, Moderna, Janssen, and AstraZeneca for use in the US and EU, there are still many individuals that will not be vaccinated due to 1) lack of access 2) ineligibility or 3) a personal choice. The worldwide vaccine roll-out itself has faced difficulties. Despite initial optimism, vaccination rates in the EU are low [26, 27]. In the US, the vaccine rollout has also faced adoption challenges. Around one third of military personnel and a similar percentage of healthcare workers have opted against vaccination, citing a variety of factors in their decision [28, 29]. Even as the vaccine is available in many parts of the U.S., several states including Minnesota, Michigan, Maryland, and Oregon have seen spikes in average new daily COVID-19 cases in the month of April 2021 [30]. Even in countries where vaccination campaigns have gone well from the standpoint of adoption rates such as Israel and the United Kingdom, severe complications due to COVID-19 are still seen [31, 32]. The emergence of novel coronavirus variants that render some vaccines less effective also threatens to prolong the pandemic. Currently there are six main variants circulating in the U.S.: the B.1.1.7 originating in the U.K.; the B.1.351 from South Africa; the P.1. first detected in Brazil; and the B.1.427 and B.1.429, both originating in California and the highly infectious B.1.617.2 (Delta) variant originating in India [33–36]. Based on decades of research on viral evolution, the emergence of other strains in the future is likely [37, 38]. As SARS-CoV-2 variants can spread faster than the original strain and some variants may be resistant to the neutralizing antibodies produced by individuals after vaccination, the risk of a widespread disease resurgence and the continued societal burdens of an extended pandemic are real [39–41].

While many molecular tests have been developed to detect the presence of the SARS-CoV-2 virus, a reduced to practice molecular test with high prognostic accuracy for severe outcomes following a SARS-CoV-2 infection represents an unmet healthcare need. Here we extended on a previously published study and describe the validation of a blood-based biomarkers for the prediction of severe symptoms of COVID-19 disease, which can place individuals in the intensive care support [15]. We identified a set of six biomarkers associated with systemic immune-genetic dysregulation, that when processed and measured by qPCR on human blood samples provided an accurate predictive readout on likely COVID-19 disease severity. While to the best of our knowledge, a comparable assay has not yet been reported on, the performance of the *EpiSwitch* assay meets the urgent need for many individuals to have *a priori* knowledge of likely disease severity in the event they are infected with the SARS-CoV-2 virus.

Interestingly, we found a significant enrichment of predictive markers at the SWAP70 gene locus. The SWAP70 locus encodes a B-cell specific ATP and calcium binding protein that is recruited to the plasma membrane following B-cell activation with subsequent translocation to the nucleus. SWAP70 has been shown to mediate immunoglobulin isotype switching in B-cells following activation of the B-cell antigen receptor complex [42, 43]. This finding is consistent with the recent reports of ongoing isotype switching in patients who are critically ill with COVID-19 and the association of differential immunoglobulin M (IgM)/IgG/IgA epitope diversity in mild or severe COVID-19, especially in patients who succumbed to SARS-CoV-2 infection [44]. We also observed that genes involved in haemostasis and blood clotting were found in the regions covered by the predictive marker panel. This is consistent with clinical reports of severe COVID-19 patients presenting clinically with a ‘microvascular injury syndrome’ with an associated procoagulant state as well as clinical reports of hypercoagulation in patients with severe COVID-19 [45, 46]. Interestingly, it remains to be investigated if the identified microvascular markers may have potential functional relevance to prothrombotic vaccination responses in individual patients. The involvement of B-cell activation and haemostasis are both in line with a prevailing view that systemic inflammation and the cardiovascular clinical manifestations it triggers, all lie at the root of the clinical symptomology seen in severe COVID-19 cases [47–49].

Extension of the current study to a wider distribution and larger number of individuals could help define the regional, racial, and epigenetic prevalence of high-risk biomarkers in these populations. A longitudinal observational study with collections before and after resolution of the acute and chronic phases of COVID disease will provide further invaluable insights into the mechanisms and the long-term stability of the identified systemic biomarker signature. Early evidence indicates that blood samples collected from patients before the onset of the COVID pandemic reveal high-risk profiles in some individuals. This would suggest that the biomarker profiles identified in this study are not emerging in response to COVID infection, but rather represent a pre-existing default state on the spectrum of outcome susceptibility.

There are several immediate implications of the results reported here. The availability of a simple blood-based assay that provides a readout of likely disease course if infected with SARS-CoV-2 is especially helpful for the triage of individuals who either 1) do not have access to COVID-19 vaccines (due to underlying medical conditions, location, or age for example) or 2) choose to forgo vaccination for other reasons. It has been well appreciated that the heterogeneity seen in COVID-19 disease outcomes are largely defined by the host response, rather than the virus or its variants [15]. The emergence of viral variants that can render some current vaccination strategies less effective and the unknown longer-term path of the virus may make information about individual risk of developing severe disease valuable for those who may be at risk of re-infection. For individuals at the early stage of a confirmed SARS-CoV-2 infection, the assay described here could assist with patient triage in hospital settings and assist in clinical decision making with a direct impact on clinical outcomes. For the currently non-infected population, the assay could help inform individuals on the potential for high-risk severe outcomes in the event of an infection. This could enable a review of preventative lifestyle decisions on vaccinations, guide appropriate social distancing measures, and inform return to work planning. Advanced knowledge of likely disease severity can also aid patients and their physicians in making preparatory medical planning decisions where appropriate. Last, when applied in larger scale, population-wide settings; knowledge of pooled individual risk profiles can help health systems make informed decisions about staffing and infrastructure needs in the event of a pandemic resurgence.

## Conclusions

Here we report on the development and validation of a predictive blood-based assay that can identify, with high accuracy, individuals who are at the highest risk of developing severe complications in COVID-19 disease. The 3D genomic biomarker panel described here can be used to evaluate personalized risk of complications prior to exposure to the SARS-CoV-2 virus. The assay described here can assist individuals and their physicians in making informed decisions on lifestyle, social, and return to work activities as well as guiding medical intervention decisions, where necessary.

## Declarations

C.K., A.W., F.S., M.S., J.W. E.H. and A.A. are full-time employees at Oxford BioDynamics plc and have no other competing financial interests.

## Author Contributions

EH, AA conceived the study, MS, EH and AR assisted with *EpiSwitch^®^* array design. WM, AB and ZL assisted with design of experiments. MS and AA planned and reviewed experiments. CK, AW, FCS and EH analysed the data. RP, AD, PB, BE, TL performed experiments. RV, AB, DP, PAR, JM helped with interpretation of the data and writing of the manuscript. WW, CK, EH, and AA. wrote and reviewed the manuscript.

## Data Availability

The data that support the findings of this study are openly available and can be found on the Github repo: https://github.com/oxfordBiodynamics/medrxiv/tree/main/CST%20publication

## Acknowledgements

The authors would like to thank members of OBD Reference Facility Morgan Thacker, Louis James, Thomas Lavin, Catriona Williams, Matt Parnell, Aemilia Katzinski, Pieter Koorts, Kalsoom Rana and Jayne Green for direct operational support and taking part in processing of clinical samples, Olly Hunter for assistance in data analysis. In addition, we acknowledge Boca Biolistics LLC. and Reprocell USA Inc. for the timely provision of high-quality clinical blood samples and clinical annotations, and Agilent Technologies, Inc. for supply of *EpiSwitch^®^* designed CGH microarray slides.

## Competing interests

The authors declare that they have no competing interests.

## Consent for publication

Written informed consent for publication was obtained from all authors.

## Availability of data and materials

The datasets used and/or analysed during the current study are available from the corresponding author on reasonable request.

## Ethics approval and consent to participate

All patients signed informed consent forms prior to providing blood samples. All ethical guidelines were followed.

## Funding

This work was funded by Oxford BioDynamics.

---

**Supplemental Table 1 & 2. Annotations and clinical characteristics for COVID-19 patient samples used in this study.** Individual characteristics and clinical features for the 116 COVID-19 patient samples used in this study (78 Training Set & 38 Test Set). CV: cardiovascular; DR: Dominican Republic; PEC: pre-existing condition; T2DM: Type 2 diabetes mellitus.

**Supplemental Table 3. Prognostic calls for high-risk with 6-marker *EpiSwitch* classifier model.** Prognostic calls of average and high risk for the 78 patients against disclosed attributes of severity: ventilation and /or ICU support.

## Notes

### Competing Interest Statement

The authors have declared no competing interest.

### Author Declarations

All clinical samples were provided by commercial suppliers Boca Biolistics LLC (FL, USA) and Reprocell USA Inc. (MD, USA), on the basis of protocol CRSPTL-00049 approved Dec 19, 2019 by Research Ethics Committee, Santo Domingo, Distrito Nacional, Dominican Republic; and Extra Institutional BB-ID-061 protocol for clinical sample collection approved by Institutional Committee of Ethics in Research, September 1, 2020, to the standards of National Health Institute of Health, pursuant to the Investigational Regional priorities and data reliability.

